# Doxycycline is a safe alternative to Hydroxychloroquine + Azithromycin to prevent clinical worsening and hospitalization in mild COVID-19 patients: An open label randomized clinical trial (DOXYCOV)

**DOI:** 10.1101/2021.07.25.21260838

**Authors:** Eugene Sobngwi, Sylvain Zemsi, Magellan Guewo-Fokeng, Jean-Claude Katte, Charles Kounfack, Liliane Mfeukeu-Kuate, Armel Zemsi, Yves Wasnyo, Antoinette Assiga Ntsama, Arnaud Ndi-Manga, Joelle Sobngwi-Tambekou, William Ngatchou, Charlotte Moussi Omgba, Jean Claude Mbanya, Pierre Ongolo Zogo, Pierre Joseph Fouda

## Abstract

**Objective:** We aimed to compare the safety and efficacy of a doxycycline-based regimen against the national standard guidelines (Hydroxychloroquine plus Azithromycin) for the treatment of mild symptomatic COVID-19.

**Methods:** We conducted an open-label, randomized, non-inferiority trial, in Cameroon comparing Doxycycline 100mg, twice daily for 7 days versus Hydroxychloroquine, 400 mg daily for 5 days and Azithromycin 500mg at day 1 and 250mg from day 2 through 5, in mild COVID-19 patients. Clinical improvement, biological parameters and adverse events were assessed. The primary outcome was the proportion of clinical cure at day 3, 10 and 30. Non-inferiority was determined by the clinical cure rate between protocols with a 20 percentage points margin.

**Results:** 194 participants underwent randomization and were treated with Doxycycline (n=97) or Hydroxychloroquine-Azithromycin (n=97). At day 3, 74/92 (80.4%) participants on Doxycycline versus 77/95 (81.1%) on Hydroxychloroquine-Azithromycin -based protocols were asymptomatic (p=0.91). At day 10, 88/92 (95.7%) participants on Doxycycline versus 93/95 (97.9%) on Hydroxychloroquine-Azithromycin were asymptomatic (p=0.44). At day 30 all participants were asymptomatic. SARS-CoV2 PCR was negative at Day 10 in 60/92 (65.2%) participants allocated to Doxycycline and 63/95 (66.3%) participants allocated to Hydroxychloroquine-Azithromycin. None of the participants were admitted for worsening of the disease after treatment initiation.

**Conclusion:** Doxycycline 100 mg twice daily for 7 days is as effective and safe as Hydroxychloroquine-Azithromycin, for preventing clinical worsening of mild symptomatic or asymptomatic COVID-19, and achieving virological suppression.

**Strengths and Limitations:**

➢ This study is one of the first randomized trial, assessing the efficacy and tolerance of Doxycycline to treat COVID-19
➢ It is one of the first to evaluate disease progression and need to hospitalization in mild or asymptomatic COVID-19
➢ Patients will not receive identical treatments
➢ Doxycycline has advantages in terms of availability, safety and cost compared to Hydroxychloroquine and Azytromycin
➢ Though this study has encounter 7 lost to follow-up, this does not have a major influence on our results
➢ These data will assist clinicians in their daily practice, and provide a new tool for the fight against COVID-19

## Background and Rationale

Since the outbreak of COVID-19, there has been no proven effective specific treatment for mild or moderate forms of disease[1]. Nonetheless, symptomatic treatments and several therapeutic agents that have demonstrated some clinical efficacy in clinical trials including remdesivir, tocilicizumab, and convalescent plasma[2]. The combination of Hydroxychloroquine and Azithromycin initially gained widespread use appearing in many national therapeutic guidelines mostly in Africa that has a past history of extensive use of chloroquine for malaria treatment and prophylaxis [1]. On the basis of *in vitro* evaluation against the severe acute respiratory syndrome coronavirus 1 (SARS-CoV-1) and SARS-CoV-2 and observational data[3], Hydroxychloroquine and Azithromycin remain the mainstay treatment of mild to moderate COVID-19 in many sub-Saharan African countries, despite concerns about possible adverse drug reactions when used in combination at high doses [4]. In the absence of approved treatment for SARS-CoV-2, many other empirical treatments are being proposed including modern and patrimonial pharmacopeia with known anti-inflammatory and anti-viral properties.

Doxycycline is a derivative of tetracycline that possesses broad antimicrobial and anti-inflammatory activities [5] [6]. Doxycycline was FDA-approved as an antibiotic in 1967 and to this day remains in the antibiotic arsenal for diverse clinical use [7]. Doxycycline has demonstrated antimicrobial, antiparasitic, and antiviral properties in several studies, including against some corona viruses[8]. Doxycyline is also generally well tolerated in clinical settings. The main objective of treating mild and moderate forms of COVID-19 is to prevent adverse progression to severe presentation requiring admission and/or intensive care. We thus aimed to compare the safety and efficacy of Doxycycline to that of the national standard guidelines (Hydroxychloroquine plus Azithromycin) for the treatment of mild symptomatic, asymptomatic COVID-19 patients receiving ambulatory care in Cameroon to prevent the disease from evolving to more severe forms potentially requiring admission.

## Methods

### Study design

The DOXYCOV trial, was an open-label, randomized, non-inferiority trial, to evaluate the safety and efficacy of Doxycycline therapy versus Cameroon National Standard therapy (Hydroxychloroquine + Azithromycin) in ambulatory patients with mild symptomatic, or asymptomatic COVID-19.

### Study site and period

The trial was conducted at the Yaounde Central Hospital Coronavirus Treatment Center between March 16 and April 09 2021. The Yaounde Central Hospital is a second category hospital in the health pyramid. It is a deconcentrated structure of the Ministry of Public Health. It has a capacity of about 650 beds for all specialties, and a considerable human resources of more than 600 employees. During the COVID-19 pandemic, it was one of the main hospital that took care of infected patients.

### Study participants

Eligible participants were 18 years of age or older, who had COVID-19 infection confirmed by SARS-CoV-2 Real-Time Polymerase Chain Reaction (RT-PCR) and were asymptomatic or with mild symptomatic with an SpO2 > 94 %. An asymptomatic patient, was define as a patient with a positive Covid-19 test with no symptoms [9]. A mild symptomatic patient, was define as a patient with a positive Covid-19 test with any of the following symptoms, including fever less than 103°F (39.4°C), fatigue, cough, sore throat, headache, muscle pain, malaise, nausea, vomiting, loss of taste and smell, lack of appetite, nasal congestion [9]. Patients were excluded if they were pregnant, breast-feeding, had currently taken trial study medication, had any contraindication to study medication, had severe cardiac disease, renal or liver insufficiency, respiratory rate ≥ 30/min, BP □ 90/60 mmHg. All participants gave written informed consent before any trial procedures were performed.

### Randomization, Allocation Concealment and Blinding

Participants were randomly assigned, in a 1:1 ratio, to receive Hydroxychloroquine + Azithromycin combination or Doxycycline combination in a central procedure performed before the start of the trial. Randomization codes were computer-generated, using the Random allocation software (Randomization Main) with a one block size. Randomly generated treatment allocation was given to clinical trial medical doctors within sealed opaque envelopes. Once a patient had consented to enter the trial, the corresponding envelope was opened and the patient was then offered the allocated treatment regimen.

### Study Procedures and Intervention

Patients assigned to Doxycycline received a fixed-dose combination of Doxycycline (100 mg PO) twice daily for 7 days. Patients assigned to the comparator arm received Hydroxychloroquine 400 mg daily for 5 days and Azithromycin 500 mg at day 1 and 250 mg from day 2 to day 5. Concomitant medications included Vitamin C 1000 mg once daily for 5 days, and Oral Zinc 20 mg once daily for 5 days in both arms.

### Data Collection and Management

Trial visits were scheduled at baseline, day 3, day 10 and day 30 where the safety of the trial drugs were assessed. RT-PCR control tests to evaluate the efficacy of the treatment were obtained at day 10. Blood samples were aseptically collected, by venipuncture of the brachial vein in a 5 ml dry tube without tourniquet. These samples were collected at baseline and day 10 for full blood count and glycaemia.

### Study Outcomes

The primary endpoint was the clinical cure rate defined as the proportion of participants becoming or remaining asymptomatic. Key secondary end points included the proportion of participants needing hospitalization due to worsening proportion, and proportion of participants with negative SARS-CoV-2 PCR test. Change from baseline in full blood count and glycaemia. All end points were assessed at Day 3, Day 10, and Day 30 in comparison to baseline. Safety evaluations included monitoring of adverse events and mortality occurring from the first dose of study treatment through the final protocol follow-up visit at Day 30.

### Sample Size and Statistical Analyses

The study was powered at 90% for the primary hypothesis tests with a 20% non-inferiority margin. Two-sided 95% confidence intervals (CI) for difference between treatments in proportion of patients with clinical cure were computed using the un-stratified method of Miettinen and Numinen [10]. For primary efficacy endpoints, non-inferiority of Doxycycline to Hydroxychloroquine-Azithromycine -based protocols was considered demonstrated if lower limit of the two-sided 95% CI for the treatment difference (Doxycycline minus Hydroxychloroquine-Azithromycin) was greater than -20 percentage points and the p value was calculated for the corresponding one-sided non-inferiority hypothesis test. Analysis of efficacy, safety and patient-reported outcomes were performed in the Intention-to-Treat population. All statistical analyses were conducted using Spss version 23.0.

### Data Safety Monitoring Board

An independent data and safety monitoring committee revised safety and efficacy data during the trial. The data and safety monitoring committee was able to pause the trial or give suggestions on potential safety issues to improve the study design and intervention.

### Patient and public involvement

Patients or the public were not involved in the design, conduct, and reporting, however the dissemination plan includes lay people summary for patients and public.

## Results

### Trial population

Of the 194 participants who underwent randomization, all received at least one dose of the assigned treatment (Fig. 1). Baseline and disease characteristics of participants are shown in Table 1. The mean age was 39 years, 47.6 % of participants were women. 42.2 % (79/187) of the analyzed participants were mild symptomatic at baseline (42/92 on the Doxycycline baseline regimen versus 37/95 on the Hydroxychloroquine-Azithromycin arm treatment, Table 1). The most common symptoms were fatigue, muscle or joint pain, anosmia, cough, headache, and chill.

**Figure 1:**
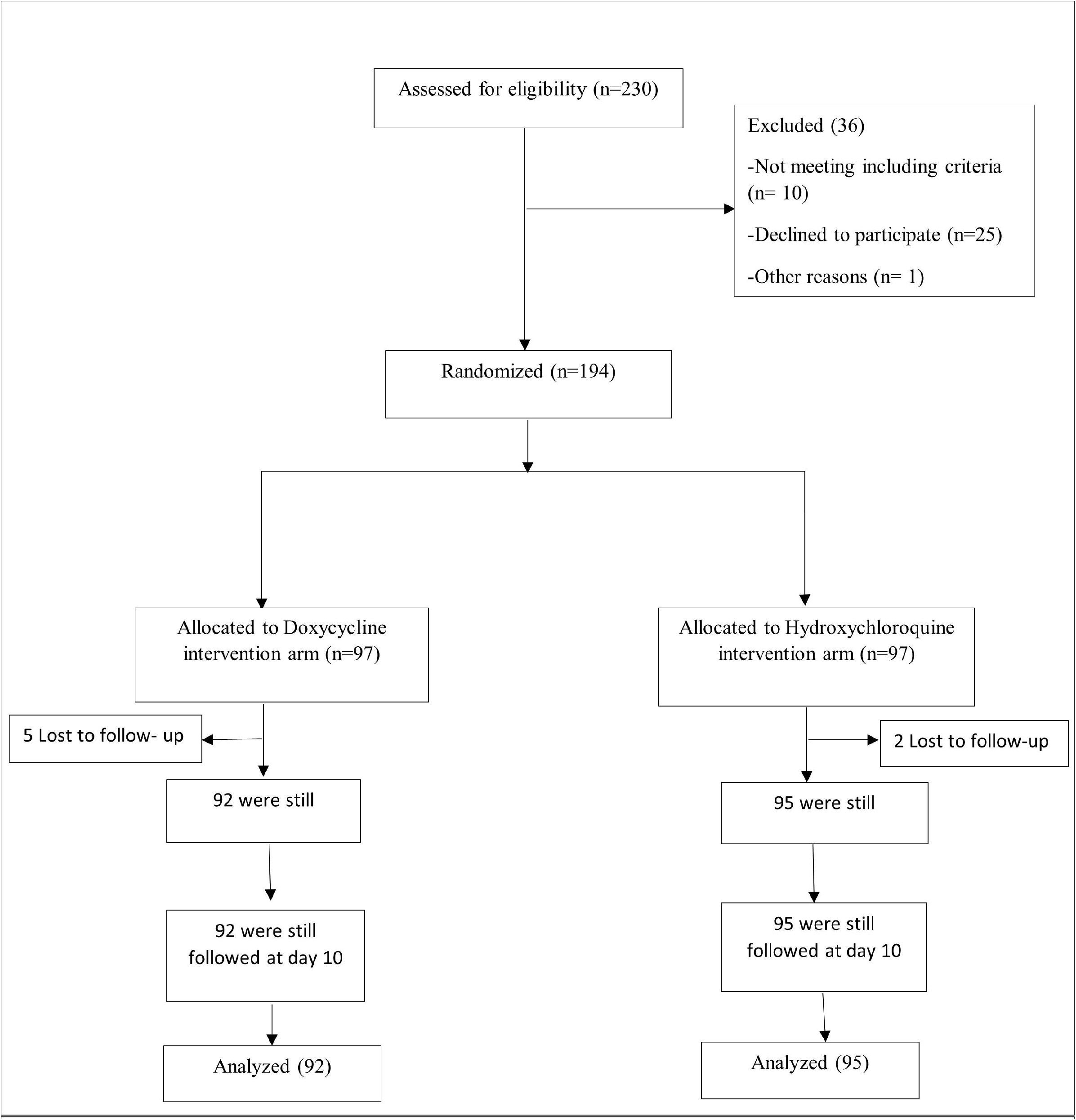
Trial profile, Intention-to-Treat Populations. Eligibility assessment, randomization and analysis

**Table 1:**
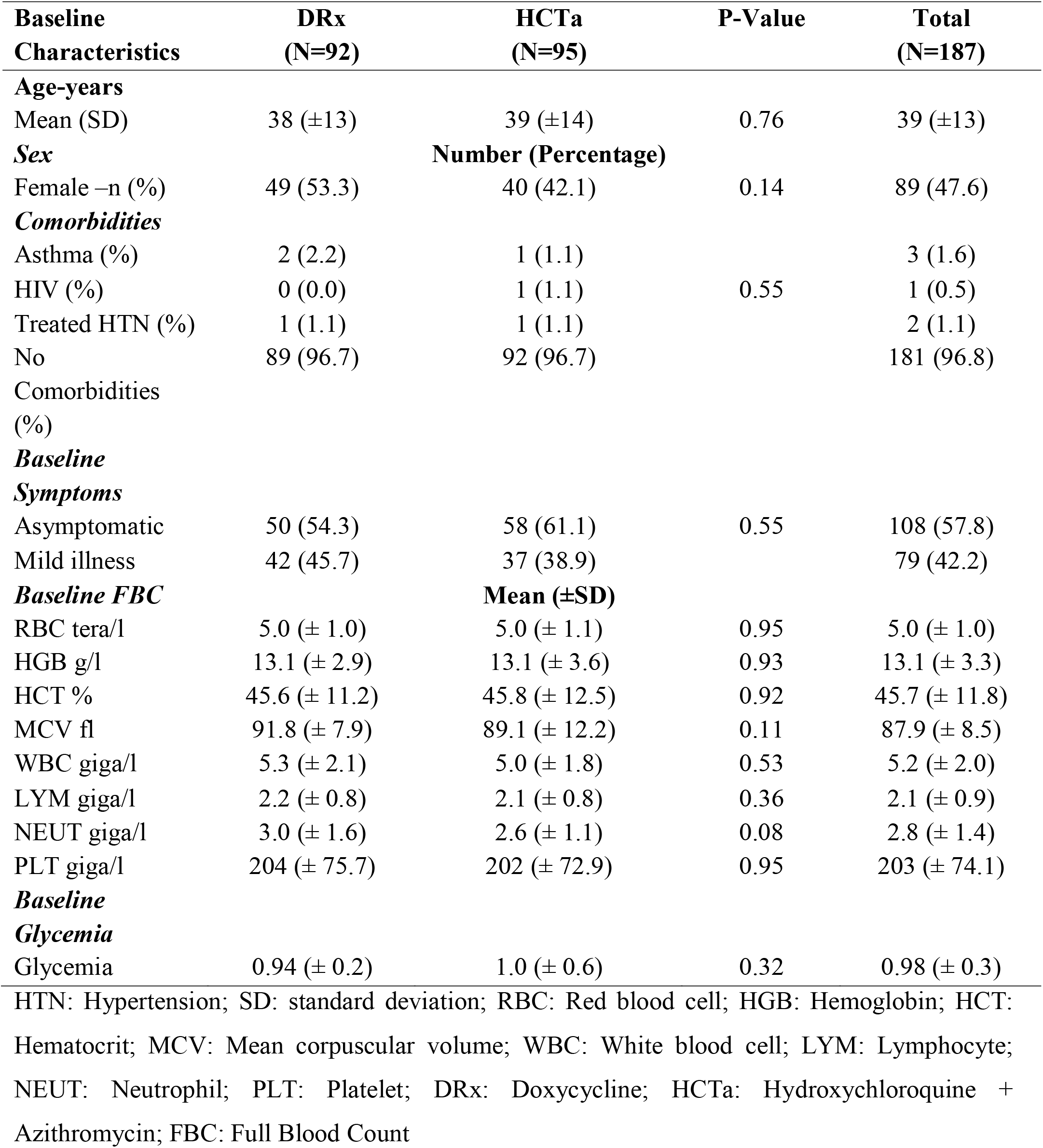
Baseline characteristics.

### Clinical and Patient-reported outcomes

Doxycycline was non inferior to Hydroxychloroquine-Azithromycin for the primary endpoint (Table 2). At day 3, the clinical cure rate was 80.4 % (74/92) for Doxycycline and 81.1 % (77/95) for Hydroxychloroquine-Azithromycin [difference (95%) CI, 0.6%(−0.1, 0.1)]. At day 10, the clinical cure rate was 95.7 % (88/92) for Doxycycline and 97.9 % (93/95) for Hydroxychloroquine-Azithromycin [difference (95%) CI, 2.2% (−0.07, 0.03)]. At day 30 all the participants were asymptomatic.

**Table 2:**
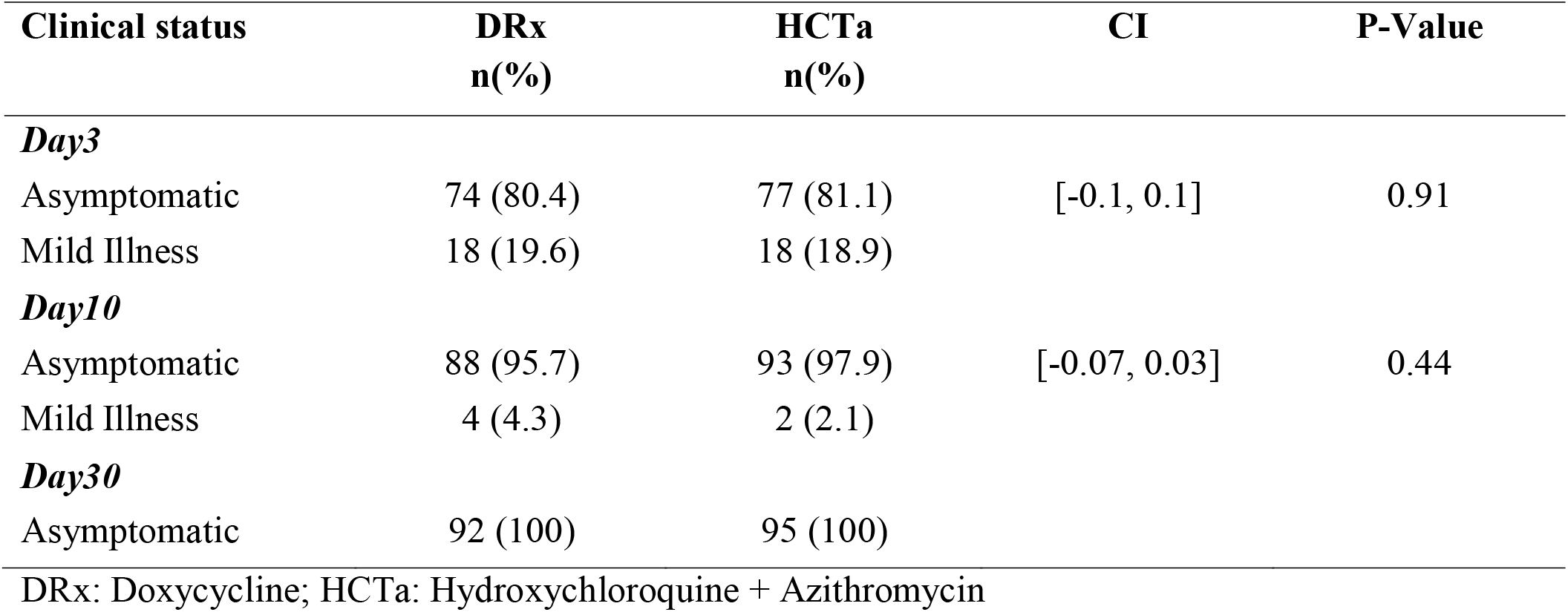
Clinical cure rate by protocol visit.

### SARS-CoV2 PCR testing

SARS-CoV2 PCR negativation rate at day 10 was 65.2 % (60/92) in the Doxycycline arm and 66.3% (63/95) in the Hydroxychloroquine + Azithromycin arm. The difference between treatment groups was 1.1 percentage points [95% confidence interval (CI), (−0.15, 0.13); p > 0.05].

### Adverse events

No major adverse event was reported. None of our participants was admitted for worsening of the disease after treatment initiation. There was no difference between treatment arms with regard to changes in full blood count and glycemia from baseline (Table 3).

**Table 3:** Biological parameters of participants at day 10.

| Biological<br>Parameters | Day 10 |  | CI | P-Value |
| --- | --- | --- | --- | --- |
|  | DRx | HCTa |  |  |
|  | Mean (SD) |  |  |  |
| RBC tera/l (SD) | 4.6 (± 0.8) | 4.8 (± 0.5) | [-0.4, 0.1] | 0.21 |
| HGB g/l (SD) | 12.2 (± 3.8) | 12.2 (± 1.7) | [-1.1, 1.2] | 0.93 |
| HCT % (SD) | 41.9 (± 6.8) | 41.3 (± 8.8) | [-2.5, 3.7] | 0.69 |
| MCV fl (SD) | 90.3 (± 7.3) | 88.9 (± 7.8) | [-1.5, 4.4] | 0.33 |
| WBC giga/l (SD) | 5.2 (± 2.4) | 5.0 (± 1.3) | [-0.6, 0.9] | 0.64 |
| LYM giga/l (SD) | 2.1 (± 0.8) | 2.2 (± 0.7) | [-0.4, 0.1] | 0.32 |
| NEUT giga/l (SD) | 3.0 (± 1.3) | 2.8 (± 1.3) | [-0.3, 0.7] | 0.46 |
| PLT giga/l (SD) | 247 (± 72.6) | 230 (± 75.8) | [-12, 46] | 0.25 |
| Glycemia (SD) | 0.99 (± 0.1) | 0.95 (± 0.2) | [-0.06, 0.15] | 0.41 |

SD: standard deviation; RBC: Red blood cell; HGB: Hemoglobin; HCT: Hematocrit; MCV: Mean corpuscular volume; WBC: White blood cell; LYM: Lymphocyte; NEUT: Neutrophil; PLT: Platelet; DRx; Doxycycline; HCTa: Hydroxychloroquine + Azithromycin

## Discussion

Since the outbreak of COVID-19 in December 2019, the world is going through waves of the pandemic of widely variable level of clinical severity. Up to 80% of people affected develop an asymptomatic or mild symptomatic disease. It remains unclear whether a specific treatment is required for mild COVID-19. However, taking into consideration the risk of adverse progression towards severe and critical forms of the disease, several therapeutic approaches are proposed worldwide to prevent need for hospitalization and admission to critical care units.

In early 2020, a small series of COVID-19 patients treated in France with HCQ showed rapid decline in SARS-CoV-2 viral load compared with controls, that seemed further improved by the addition of Azithromycin [11]. However, methodological flaws were reported, limiting the interpretation of the results [12], in addition to few other conflicting trials on the use of Hydroxychloroquine, and its possible cardiac adverse events [3]. Nevertheless, some countries especially in sub Saharan Africa still recommend HCQ-Azithromycin as the reference treatment in the management of COVID-19 patients [13]. The development of vaccines is expected to drastically reduce hospital and intensive care admissions as well as death. In the meantime, many other treatment options for mild and moderate COVID-19 were investigated to mitigate progression towards severe disease, hospitalization or death with variable efficacy, including mostly drugs with antiviral and anti-inflammatory effect [14,15].

In this randomized, open-label trial, we compared Doxycycline with Hydroxychloroquine-Azithromycin for the treatment of ambulatory patients with mild symptomatic or asymptomatic COVID-19. None of the participants progressed to severe disease nor required hospital admission, and we showed the non-inferiority of a 7-day course of Doxycycline to a 5-day course of Hydroxychloroquine-Azithromycin in terms of clinical cure rate, virological suppression, and safety profile. No major adverse event was recorded possibly due to the young age of the study population, the safe dosage used in our participant (400 mg Hydroxychloroquine and 250 mg Azithromycin), and prior exposures to the study drugs for malaria, and in accordance with our preliminary safety study [16].

Our results support the observation by Mohammud M Alam et al. in 2021 in the United States of America, where they evaluated the clinical outcomes of early treatment with doxycycline for 89 high-risk covid-19 patients in long term care facilities, and found that early treatment with Doxycycline for high-risk patients with moderate to severe COVID-19 infections in non-hospital settings, is associated with early clinical recovery, decreased hospitalization, and decreased mortality [17]. The present trial was not placebo-controlled, but the >95% cure rate at Day 10 contrasts with 70% reported in placebo groups at Day 14 in other trial [18].

While the study population was relatively young with limited comorbidities, it reflects the general population of Cameroon and most sub-Saharan countries. In 2019, the Cameroon Demographic Health Survey, estimated that, 60% of the population is aged 20 to 40 years old, and only 14% of the population is above 40 [19]. In the earlier stages of the COVID-19 pandemic, older age was considered among important factors associated with clinical severity [20], however, recent reports indicated for example in more than 3000 adults ages 18 to 34 who contracted COVID-19 and became sick enough to require hospital care, 21% ended up in intensive care, 10% were placed on a breathing machine and 2.7% died [21]. Young adults are therefore a major target population for interventions that would reduce progression to severity to mitigate the public health and economic effects of the pandemic.

In conclusion, despite a modest 65% PCR negativity at Day 10, we report a remarkably high clinical cure rate in response to a Doxycycline based treatment protocol for mild COVID-19 with a safety profile that supports its use as an alternative to prevent adverse disease progression and need for hospitalization.

## Data Availability

Data availability is subject to prior approval by the National Ethics Committee of Cameroon and the Minister of Public Health

## Acknowledgments

The authors greatly thank all the personnel of the COVID-19 Unit at the Yaounde Central Hospital. The authors are also grateful to all participants who have voluntarily accepted to be enrolled in this study.

## Patient consent

Obtained

## Data confidentiality

The authors declare having followed the protocols in use at their working center regarding patient’s data publication.

## Ethical approval

The study received ethical clearance from the Cameroon National Ethics Committee (N° 2020/07/1585/L/CNERSH/SP). The trial was prospectively registered on ClinicalTrials.gov under number NCT04715295.

## Data Statement Section

Due to the sensitive nature of the questions asked in this study, our participants were assured that data would remain confidential and would not be shared.

## Author Contribution Statement

Contributors: E-S designed the work, acquired funding, implemented the trial, monitored data collection for the whole trial, monitored statistical analysis, drafted the paper, revised the paper and gave final approval to be published. S-Z designed the work, implemented the trial, monitored data collection, wrote the statistical analysis plan, cleaned and analyzed the data, drafted and revised the paper. M-GF designed the work, revised the draft paper. JC-K designed the work, revised the draft paper. CK member of the data safety monitoring board. LM-K member of the data safety monitoring board. AZ designed the work. YW designed the work, collected data. AA-N collected data. AN-M collected data. JS-T monitored data collection, monitored statistical analysis, revised the paper. WN member of the data safety monitoring board, revised the paper. CMO member of the data safety monitoring board. PJF member of the data safety monitoring board. POZ member of the data safety monitoring board. JCM member of the data safety monitoring board, revised the paper.

